# Perennial Disaster Patterns in Central Europe since 2000: Implications for Hospital Preparedness Planning – A cross sectional Analysis

**DOI:** 10.1101/2024.03.10.24304050

**Authors:** M von der Forst, M Dietrich, FCF Schmitt, E Popp, M Ries

**Affiliations:** Heidelberg University, Medical Faculty Heidelberg, Department of Anesthesiology, Im Neuenheimer Feld 420, 69120 Heidelberg; Heidelberg University, Medical Faculty Heidelberg, Pediatric Neurology and Metabolic Medicine, Center for Pediatrics and Adolescent Medicine, Im Neuenheimer Feld 430, 69120 Heidelberg

**Author notes:** **Corresponding Author:** Dr. med. Maik von der Forst, Im Neuenheimer Feld 420, 69120 Heidelberg.

**Keywords:** Disaster Management, Mass Casualty Incident, Hospital Emergency Preparedness, Natural Hazard, Hospital Disaster Planning

## Abstract

**Introduction:** Hospitals are vital components of a country’s critical infrastructure, essential for maintaining resilient public services. Emergency planning for hospitals is crucial to ensure their functionality under special circumstances. The impact of climate change and seasonal variations in the utilization of hospital services further complicate emergency planning. Therefore, the knowledge of perennial disaster patterns could help strengthening the resilience of health care facilities.

**Methods:** We conducted a cross-sectional analysis of the Emergency Events Database EM-DAT for disasters in Central Europe between January 2000 and December 2023 (defined as Germany and bordering countries). Primary endpoint was the average month of occurrence of disasters across the overall study period.

**Results:** Out of 474 events, 83% were associated with a natural cause and only 80 events (=17%) were technological. More than 50 % of the technological disasters were categorized in the transport accident subgroup. Technological disasters were spread equally over the whole year. The vast majority of natural disasters (N=394) were due to storm (n=178, 45%), flood (n=101, 26%) and extreme temperatures (n=93, 24%) with peaks occurring during summer and winter months, while less disasters were registered during autumn and especially spring seasons.

**Discussion:** Looking at the three most common disaster types, extreme temperatures, floods, and storms are clearly dominating and cause over 90% of the natural disasters in central Europe. An overlap of hospital admissions due to seasonal effects and catastrophic events, mainly triggered by natural disasters in the vulnerable periods may lead to a partial collapse of the health care system. To deal with such a variety of different and potentially simultaneous hazards using an “all hazards” approach could be promising and often has been seen as the most effective strategy for hospital emergency planning.

## Introduction

Hospitals are one of the main institutions of the health care system. As a part of the so-called critical infrastructure of a country they are essential for the maintenance of public services (1). Hospital emergency planning is crucial to ensure that their functionality can be maintained also under special circumstances. Strengthening the resilience of the critical infrastructure and among them health facilities is one of the goals of the Sendai Framework for disaster risk reduction until 2030 (2) In different countries hospitals are obligated to prepare plans for different potential emergency situations e.g. mass casualty events, fire, or blackout. There are existing national and international guidelines and reviews which define the most important scenarios to prepare for (3–7). Most of the mentioned threats handle local problems and assume an intact infrastructure as well as the possibility of regional assistance through external authorities. As such, a survey of 96 German hospitals showed that only about 25% of the hospitals have plans for floods or extreme weather events (8). But in the last years in Europe there were a number of events at a larger scale, like the “Ahrtal Flood”, different wildfires or heatwaves which compromised more than a single region at the same time and led at least partially to a destroyed or compromised infrastructure. Similar phenomena were also seen after the earthquake in Algeria 2003 and the Indian Ocean tsunami of 2004 which led to a significant reduction of health facilities (9).

In addition to these catastrophic events partially due to climate change, the utilization of hospital services underlies seasonal differences. On the one hand the summer months could be associated with a higher number of heat-associated illness e.g. cardiovascular and renal diseases on the other hand during the winter season respiratory virus infections are common (10–12). It is therefore essential to have an overview about the origin and the perennial pattern of disasters to prepare hospitals for these future challenges and help hospital administrators and emergency managers to prioritize resources. While there are many case reports and analysis for smaller events affecting hospitals (fire, traffic incidents, terroristic attacks, cybercrime), for Europe at our knowledge at the moment there is no analysis which kind of events at the disaster level may predominantly affect hospitals (13–17).

We asked the following research question:

Across the study period 2000-2023, were there particular months or seasons where disasters had particularly peaked, in the overall average, and which disasters were these?

With the present cross-sectional study of the EM-DAT database we aim to analyse and characterize disaster types in central Europe since 2000 with a particular focus on perennial disaster patterns and outline the potential impact of these findings for future hospital emergency planning and better preparedness.

## Methods

We conducted a cross-sectional analysis on disaster events the Emergency Events Database EM-DAT (18). EM-DAT is a free open access disaster database, considering disasters from all over the world (19). We included listed events between January 2000 and December 2023 in Central Europe, which was defined as Germany and bordering countries (Denmark, The Netherlands, Belgium, Luxembourg, Switzerland, Austria, Czech Republic, and Poland). Events before 2000 were not taken into consideration because “*Pre-2000 data is particularly subject to reporting biases*”, further we considered this time period of 23 years as the most current and best representative sample for the research question (18). The Covid-19 pandemic as an exceptional and still ongoing event affecting people worldwide was excluded from the analysis, because it has special emergency implications due to its magnitude and is considered different from the usual underlying disaster pattern. The selection of countries was based on two particular assumptions: 1) there should be a geographical proximity to make climate variables and exposure to the same kind of events comparable, 2) the socio-economic status of a country and technological development should be similar due to their similar implications on necessary hospital preparedness.

All listed disaster events were defined as situations which: *“overwhelm local capacity, necessitating a request to the national or international level for external assistance; an unforeseen and often sudden event that causes great damage, destruction, and human suffering*.*”* (18). To be listed an event has to further met at least one of the following EM-DAT Inclusion Criteria: at least ten deaths (including dead and missing), or at least 100 affected (people affected, injured, or homeless) or a call for international assistance or an emergency declaration (18). Primary data sources for EM-DAT include “UN agencies, non-governmental organizations, reinsurance companies, research institutes, and press agencies” (18).

Disasters are listed with four levels of depth, so events are divided into groups, subgroups, types, and subtypes. The three EM-DAT disaster groups are ‘Natural’, ‘Technological’, and ‘Complex’. The Peril Classification and Hazard Glossary is currently the primary reference for classifying natural hazards and divides them into six main subgroups: Geophysical, Hydrological, Meteorological, Climatological, Biological, and Extra-terrestrial (20).

Data were downloaded from the EM-DAT website on 12^th^ December 2023 and considered events including November 2023, and then exported into an electronic database system (Microsoft Excel®, Microsoft Deutschland GmbH, Unterschleißheim, Germany). Variables for analysis included: [Disaster Group, Disaster Subgroup, Disaster Type, Country, Start Year, Start Month, Start Day, End Year, End Month, End Day, Total Deaths]. The statistical analysis and figures were performed with SPSS (Statistical Product and Services Solutions, Version 25, SPSS Inc., Chicago, IL, USA) and Graphpad Prism (Version V, GraphPad Software, La Jolla, USA). Standard methods of descriptive statistics were applied in the analysis of the complete dataset.

Primary endpoint was month of occurrence across the overall study period. We compared the disaster groups natural and technological. The perennial pattern of disasters was elaborated by attributing all disasters across the overall analysis period into their individual month of occurrence, irrespective of the year. Further the disasters were divided into types following the EM-DAT nomenclature, the perennial pattern of types was analysed in the same matter. Every disaster event was listed by the country which was affected and then compared to the ground area and the number of inhabitants of every country.

The analysed years were further divided into two equal time periods to compare the frequency of disaster during a longer period. Also, the duration in months of every event was analysed.

For a comparison of disaster number and the total Inhabitants of the different countries the German Website Statista.de was used (21) The study fulfills the requirements of the STROBE statement (22).

Ethics: As a descriptive analysis based on pre-existing data without direct human or animal research involvement, the present study did not require Institutional Review Board Ethics Committee review.

## Results

Since 1^st^ January 2000, 474 events were registered in EM-DAT. Fifty-four percent of all disasters occurred during the first half of the mentioned time period. Overall, the number of disasters in central Europe, as defined for the underlying study, was slightly decreasing over time.

The number of events categorized by month of occurrence across the overall analysis period shows that the disaster peaks are situated during summer and winter months, while less disasters are registered during autumn and especially spring seasons. Compared to technological events the frequency of disasters related to natural phenomena was much higher, since 2000 there were registered 394 events in EM-DAT. As a characteristic property of disasters due to natural hazards that were included in this analysis, a varying frequency during the course of the year with a particular pattern could be detected (Fig. 1). Going more into detail reveals that these periodical fluctuations can be associated predominantly to the three main types of natural disasters, i.e., extreme temperatures, flood, and storms (Fig. 1).

**Figure 1:**
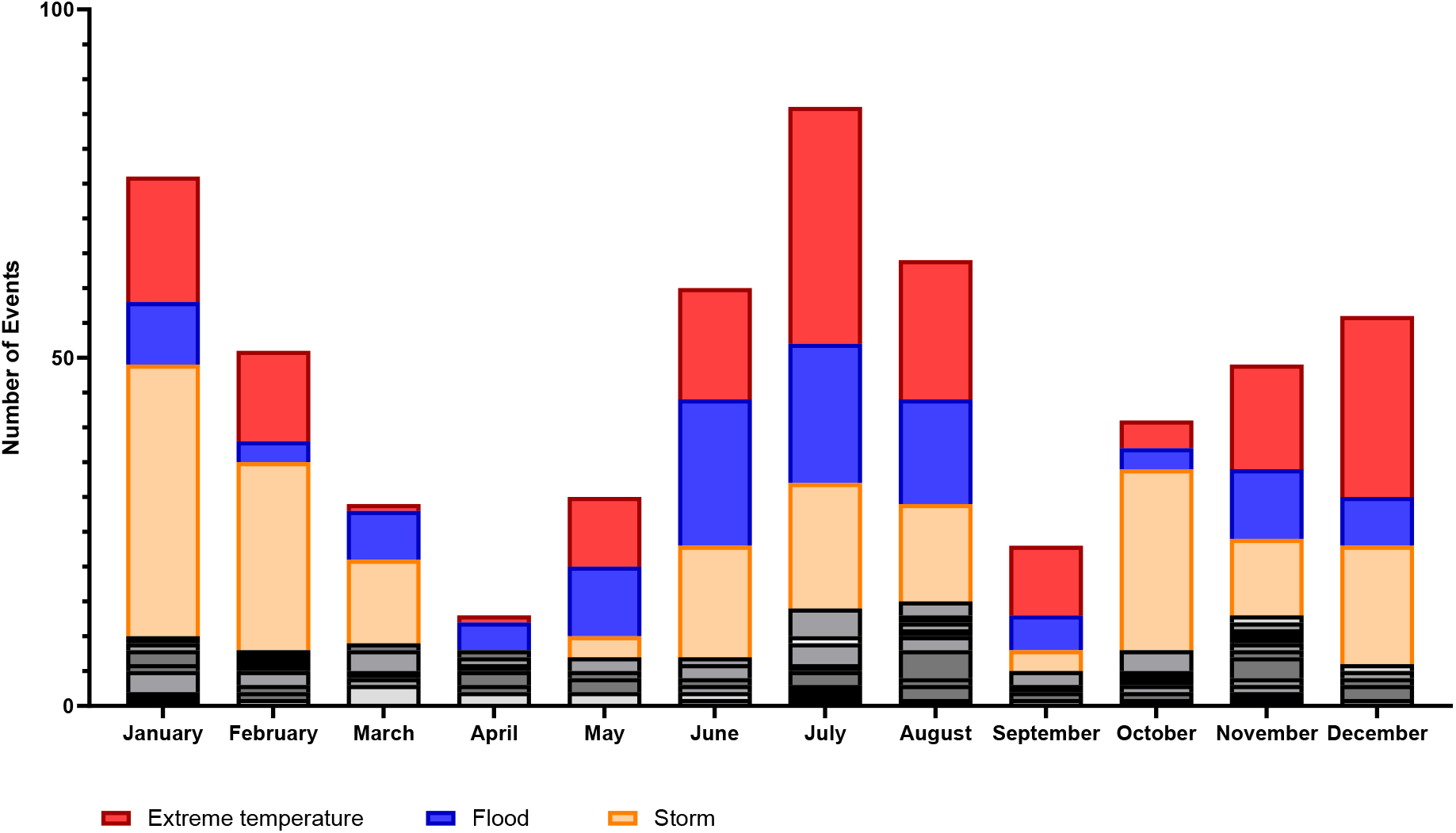
Perennial pattern of disasters in Central Europe by month of occurrence between January 2000 and November 2023 (in black all other disaster types).

While the were some minor causes like drought (n=2), earthquake (n=3), epidemic (n=4), mass movement (n=8) and wildfire (n=5), which taken together represent only 5.6% of all listed natural disasters, the vast majority of disasters were due to storm (n=178, 45%), flood (n=101, 26%) and extreme temperatures (n=93, 24%).

It appears that while storms are most frequent in the winter season, more floods are registered during summer months (Fig. 2). For extreme temperatures there are two peaks one for extreme heat und one for extreme cold, corresponding to summer and winter months (Fig. 2). Importantly, while the absolute number of cold events is higher (n= 52 vs. 41) than extreme heat, the impact regarding death is nearly completely associated to heat events (97% of all extreme temperature deaths). Considering all events there were 60 which had a duration > 1 month from start to end, 55 of these belong to the three dominating disaster types storm, flood and extreme temperature. The maximum duration of a disaster was 5 months (extreme temperature). In general, extreme temperature disasters lasted longer than floods and storms.

**Figure 2:**
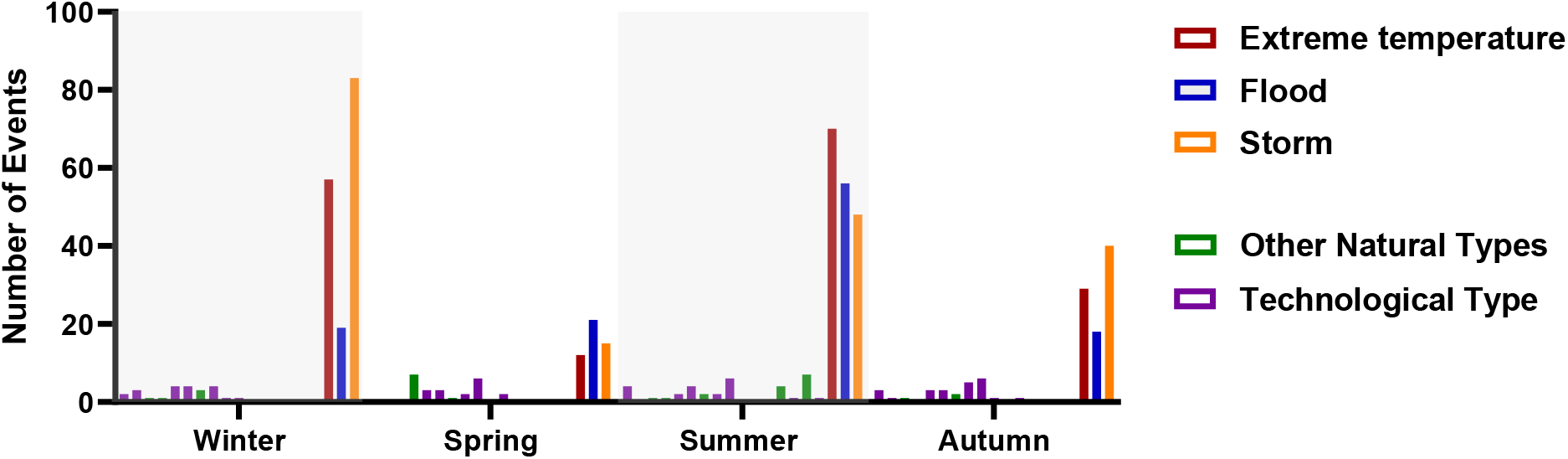
Overall number of disasters by type in Central Europe between 2000 to 2023, categorized by season of the year (in black all other disaster types).

A better visualisation of the three most common hazards during the course of the year and the analysis of potential coincidences is given by a heat map which is a graphical representation of data where values in a matrix are represented as colors (Fig. 3). The numerical magnitude of events is represented by colours whereas light green means zero events and red means maximum number of events (in our dataset fourty). The Months March, April and May show the lowest incidence of the main disaster types in central Europe, further also September, October and November from 2000-2023 were less affected by disasters than Winter or Summer Months (Fig. 3). Some major characteristics of the three main disaster types are shown in Table 1.

**Table 1:**
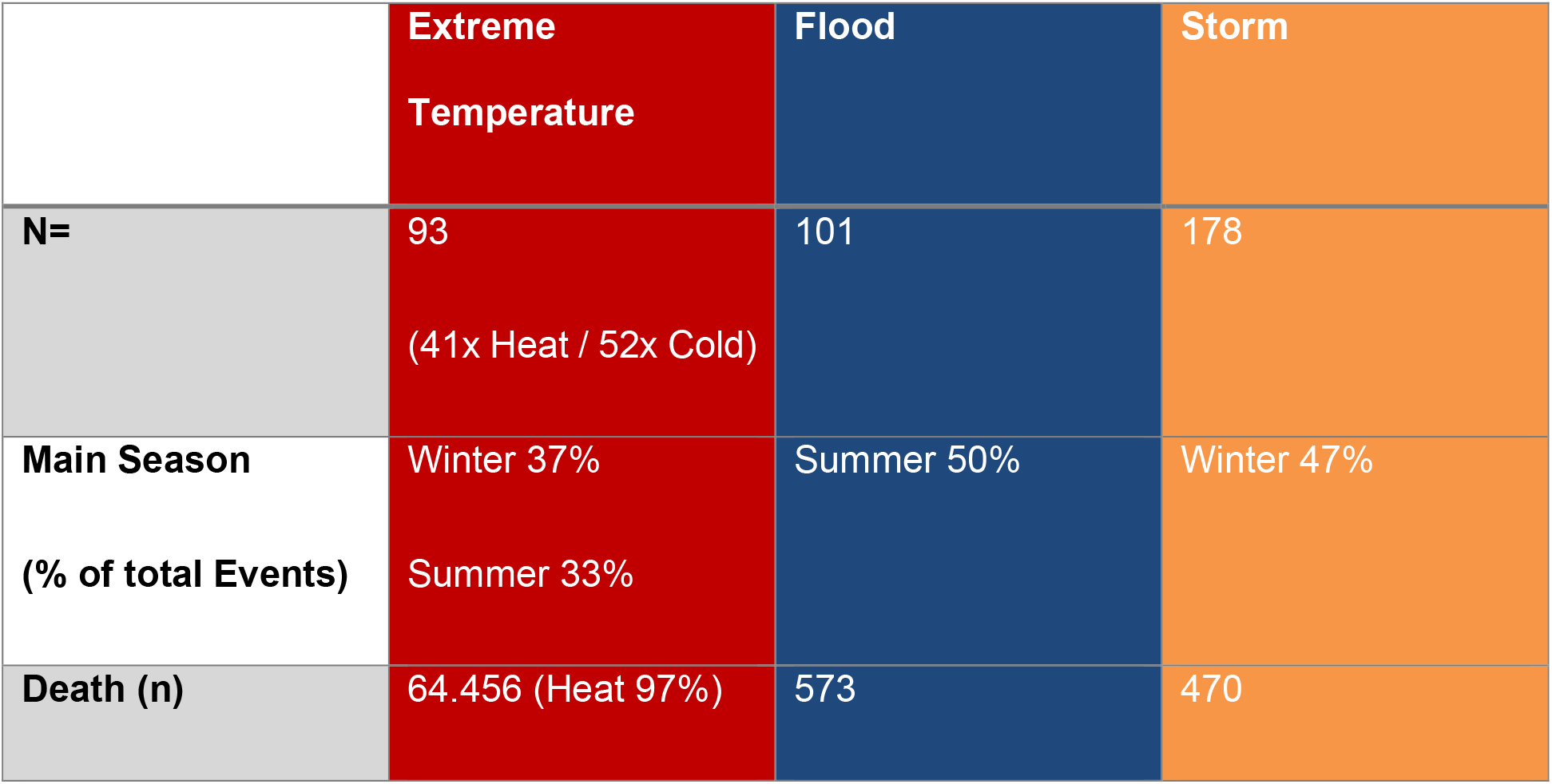
Main season and deaths associated with disasters due to extreme temperature, flood, and storm in Central Europe between 2000 and 2023.

|  | Extreme Temperature | Flood | Storm |
| --- | --- | --- | --- |
| N= | 93<br>(41x Heat / 52x Cold) | 101 | 178 |
| Main Season<br>(% of total Events) | Winter 37%<br>Summer 33% | Summer 50% | Winter 47% |
| Death (n) | 64.456 (Heat 97%) | 573 | 470 |

**Figure 3:**
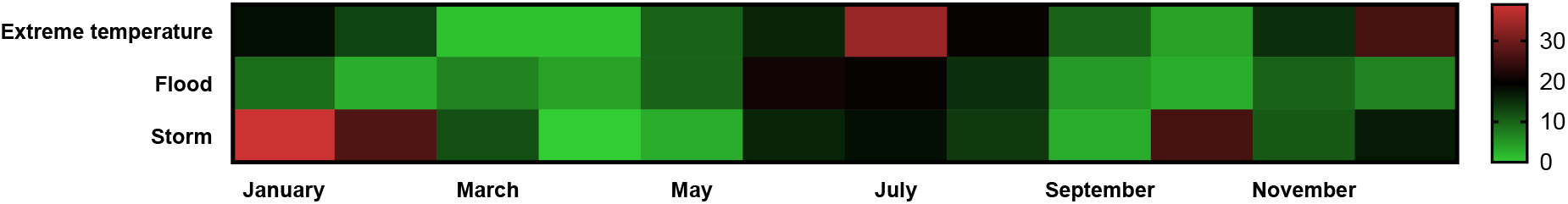
Heat map showing perennial frequency distributions for extreme temperatures, flood, and storm in Central Europe between 2000 and 2023.

In total most of the analysed disasters were associated to a natural cause and only 80 events (=17%) were technological. More than 50% of the technological disasters were categorized in the transport accident subgroup (aircraft n=9, rail n=13, road n=19, water n=2). The remaining events (n=37) were all of the subgroup industrial and miscellaneous accidents. In detail there were mostly fires (n=14) or explosions (n=13) or collapses (n=4). There were no substantial differences between the analyzed countries, whereas, of interest, the bigger industrialized ones in total had more technological events (e.g. France n=23, Germany n=19). Looking at the different months, there was a slight fluctuation for technological disasters over the whole year. The months January, July and November have more events in total, but these could not be attributed to specific causes and seems coincidental (Fig. 4). Dividing the analyzed period into equal parts (2000-2011 & 2012-2023) it appears that technological events are becoming less frequent and 70% (n=56) of the registered disasters happened between 2000 and 2011.

**Figure 4:**
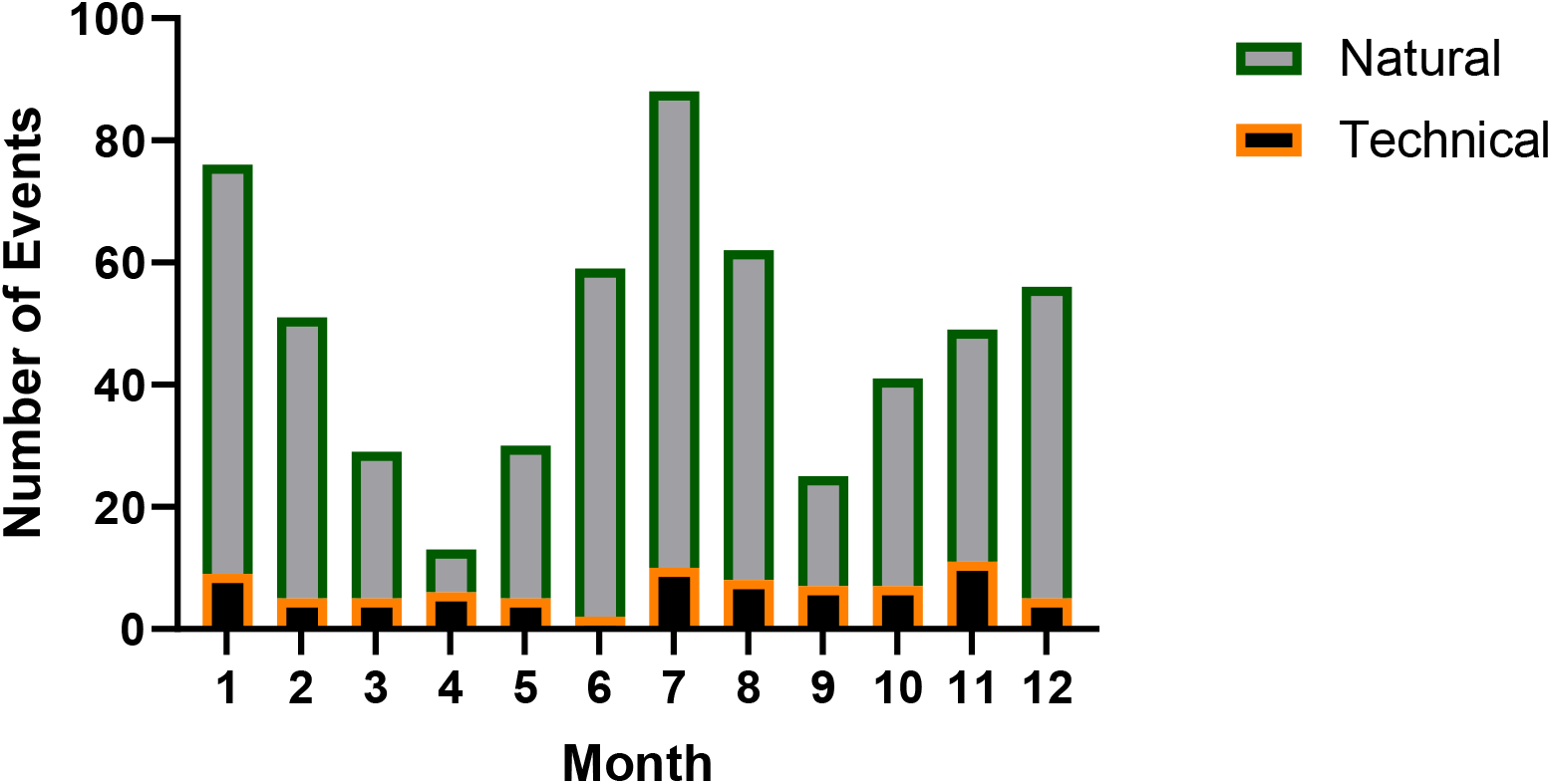
Number of disasters in Central Europe between 2000 and 2023 attributed to month of the year across the overall study period, stratified by natural or technical types.

As expected, the number of disasters was different between the selected countries. France (n=137) and Germany (n=83) showed the highest total number of disasters. Comparing these data with the area and the population size of the countries it results that both France (2.1 Events/Mio. inhabitants) and Germany (0.96 Events/Mio. inhabitants) are together with Denmark (1,7 Events/Mio. Inhabitants) the three countries with a relatively smaller number of disasters, while e.g. in Luxembourg there were 9,2 Events/Mio. inhabitants (21).

The three main disaster types correspond to the most common in almost all analyzed countries. There are only a few exceptions regarding flood events. Denmark (0% of all events), the Netherlands (4% of all events) and Switzerland (10% of all events) had relatively few flood disasters registered in the database (Fig. 5).

**Figure 5:**
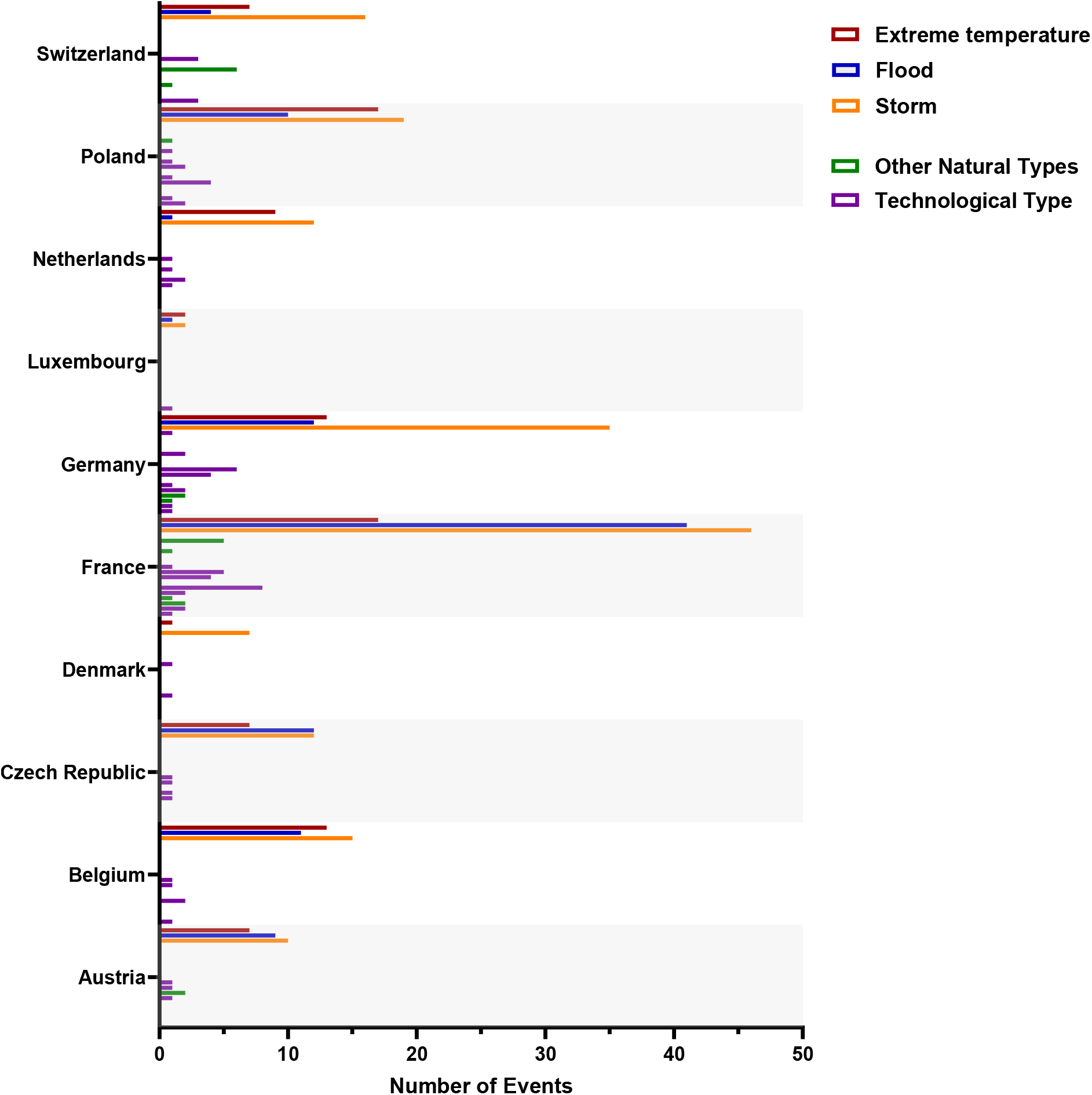
Absolute number of disasters between 2020 and 2023 categorized by country and disaster type (in black all other disaster types).

Comparing the first and the second half of the analyzed time period shows that the number of storms and extreme temperature events is rising, while the number of registered floods is numerically decreasing as well as the sum of technological incidents (Fig. 6).

**Figure 6:**
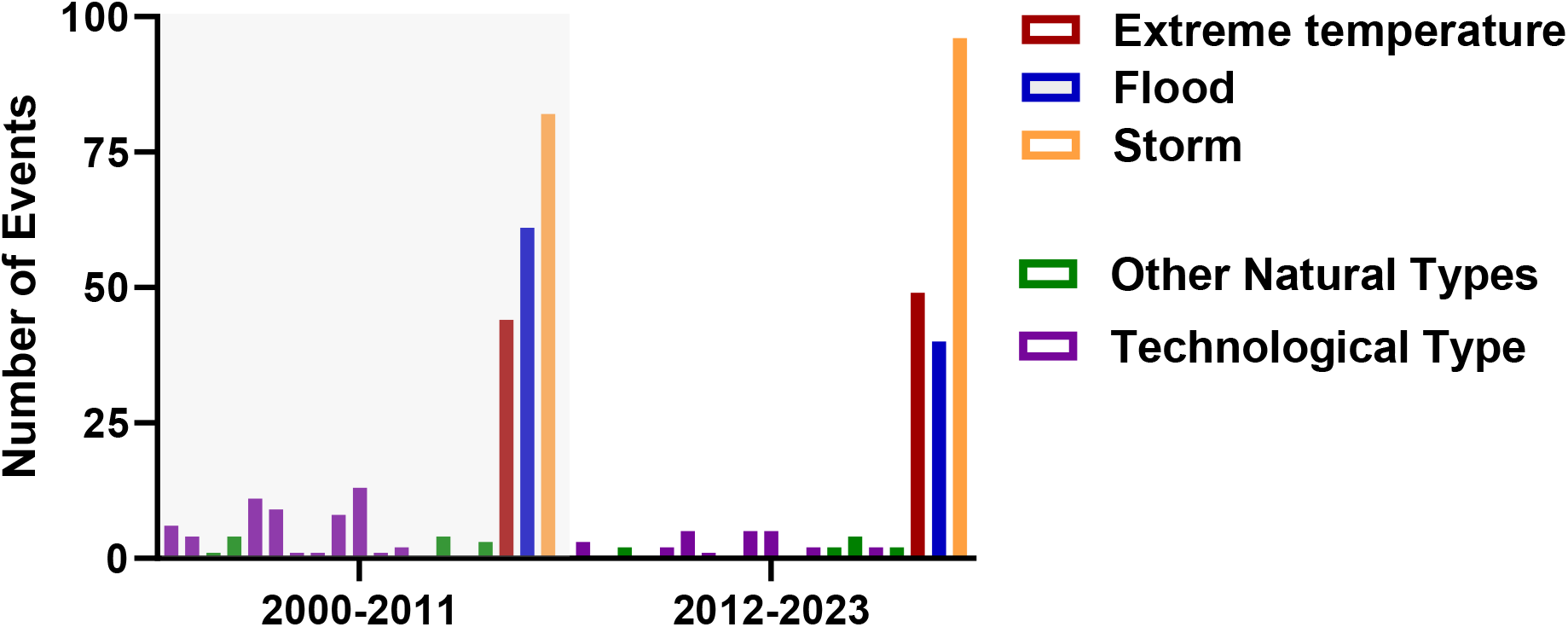
Overall number of disasters 2000 – 2023 in Central Europe categorized by semester of observation period and disaster type (in black all other disaster types).

## Discussion

The present analysis points out the perennial pattern of disasters across central Europe between 2000 and 2023 and highlights critical peak seasons necessitating hospital preparedness. Notably, our examination reveals that the summer and winter months emerge as the most vulnerable periods, characterized by a prevalence of extreme temperatures, floods, and storms. Particularly alarming is the disproportionate loss of life attributed to heatwaves during the summer months. While other regions of the globe have grappled with the impact of natural hazards such as floods, hurricanes, and droughts on healthcare infrastructure for decades, Central Europe has, fortunately, been largely spared by such events in recent years (23–27). Preparation and precaution are nevertheless essential to diminish the number of deaths in these potentially upcoming disasters (28). An essential factor is that public healthcare could be affected much longer than the event itself (29). Different events of the last few years e.g. the “Ahrtal Flood” confirmed this perception, some hospitals that were hit by the flood and in part had to be evacuated, were not able to get back to their usual business for several months (30–32). Unfortunately, the level of hospital preparedness is varying and probably due to the lack in legislation or financial issues it is often insufficient (33). Especially in industrialized countries compared to developing countries the focus of disaster planning lies less on natural hazards (34).

The analysis of registered catastrophes in central Europe during the last 23 years showed that the main group are natural hazards (Fig. 1) which affect the different countries in a similar manner (Fig. 3). The results of the ongoing analysis further indicate that especially during summer and winter months (Fig. 2), the incidence of catastrophic events is higher than in spring and autumn. Both are seasons where the demand of health services usually is already high and hospitals are often working at their limits e.g. due to heat or the annual epidemics of different respiratory viral diseases (10). Looking at the three most common disaster types extreme temperatures, floods and storms are clearly dominating and cause over 90% of the disasters related to natural hazards in central Europe (Fig. 2). In the hospital disaster planning literature, a publication form Munasinghe et al. which analyzed 53 publications, less than 10% and in a systematic review by Hasan et al. only two of thirteen articles were dealing exclusively with these three hazards (34,35).

Taken together extreme temperatures appear during the summer and the winter season, whereas especially heat periods are a major health problem and cause a significant number of deaths (36–38). This is mostly due to heat-related illness including e.g. cardiovascular and respiratory complications as well as renal failure and negative impact on fetal health (11,39). The results of the distribution further show that floods have a peak during summer season while, vice versa, storms appear predominantly during late autumn and winter months. This further means a probability of a coincidence of two disaster types at the same time (e.g. heat wave and flood) (Fig. 6). Further especially these leading hazards in the future will probably become more frequent and affect more people in different regions due to climate change (25). An overlap of hospital admissions due to seasonal effects and catastrophic events may lead to a partial collapse of the health care system.

Looking at the results of the present study and the findings of the literature it should be of high interest and priority to prepare hospitals for potentially overlapping disasters. Financial issues should not be an obstacle as in the U.S. weather and climate disasters since 1980 caused a total damage that exceeds $2.655 trillion (40). In total compared to other topics there is still only few scientific literature regarding catastrophic events and hospital emergency planning, also if there is a rising number of scientific articles coming up especially after larger disaster events (41). An analysis from Melnychuk et. al found 363 peer-reviewed publications including the grey literature. Sorted by type it shows that there are 13 different kinds of threats and over >45% of the peer reviewed literature belong to the category “Others” (42). Other reviews further indicate that in the majority of hospitals some crucial aspects of emergency planning are missing, these include especially dead body handling, waste management, transport and access routes, which are relevant mostly in disaster situations with a destroyed infrastructure (34). A systematic review by Hasan et al. had similar findings, screening over 2500 publications led to 13 peer reviewed articles focusing on hospital surge capacity preparedness. Overall missing stuff and staff, the lack of standardized assessment tools and system related barriers were identified as the limiting factors to preparedness for surge capacity (35).

Taken together because of the complexity of this topic hospitals should engage experts which develop a hospital disaster plan/ business continuity plan (often used synonymously) (4,43). To deal with such a variety of different and potentially simultaneous hazards using an “all hazards” approach could be promising and often has been seen as the most effective strategy for hospital emergency planning (44,45). The findings of two different reviews demonstrate that the all-hazard approach is an established strategy and was chosen in more than half of the articles (34,35). Preparing for “all-hazards” means that the processes of emergency planning are organized without knowledge of the specific event. On the one hand this model has some advantages especially in case of disasters that previously were not considered in the plans of a single hospital on the other hand it could also have potential negative effects regarding the preparation of hazard specific tasks (46). A mixture of both strategies e.g. by creating toolbox including hazard specific protocols but using an “all-hazard” approach as a backbone for command and control, communication and alert issues could strengthen the resilience of health care facilities. The main task to fulfill the goals of the Sendai Framework should be the development of constructive, infrastructural, and administrative resilience (2). To operationalize disaster preparedness Verheul et. al developed nonagon with crucial components (47). It could be helpful to consider the 2015 updated Hospital Safety Index tool, developed by the World Health Organization and the Pan American Health Organization, which is the mostly used tool for hospital emergency preparedness assessment (48).

This analysis relies on the accuracy of the data collection in EM-DAT. The database driven design of pre-existing data with unchangeable elements is a limitation regarding the possibility of a more specific analysis. It cannot be excluded that recording of disaster events may be incomplete (49). Nevertheless, during the analysis period covered in this report, data acquisition into EM-DAT was performed prospectively. Further in some of the listed events there are missing data, especially regarding the impact (costs, death, affected people), but this was not the main focus of the actual analysis. In general, EM-DAT is considered an authoritative data source in the field. Another relevant limitation is that some disasters which affected more than one country at the same time were listed as a single entry in EM-DAT for every country and the given data did not offer the possibility to group these events reliably. Therefore, the same catastrophe appears once for every single affected country, which may lead to an overestimation. On the other hand, the advantage of this method is that events hitting larger geographical areas appear with a higher impact by their higher single number.

## Conclusion

The awareness of perennial disaster patterns in central Europe with particular vulnerability phases in the summer and winter months may allow hospital managers and emergency planners better preparation, response, and recovery from predominant disaster events, mainly extreme temperatures, flood, and storm. In other parts of the world the high frequency of climate associated disaster events and the analysis of their potential damages as well as community disruptions already led to the development of toolkits to prepare the health care system for changing climate(50,51). Taken together potential upcoming of more than one scenario at the same time should be considered, therefore a mixed approach of event specific protocols and an “all hazard” strategy for core areas could strengthen the adaptive resilience of health care facilities.

## Data Availability

https://www.emdat.be/

## Notes

### Competing Interest Statement

The authors have declared no competing interest.

### Funding Statement

This study did not receive any funding

